# Monogenic Parkinson’s Disease and the Impact of *APOE* E4: A Case-Control Study

**DOI:** 10.1101/2024.11.15.24317402

**Authors:** Matthew J. Kmiecik, Michael V. Holmes, Pierre Fontanillas, Giulietta M. Riboldi, Ruth B. Schneider, Jingchunzi Shi, Anna Guan, Susana Tat, Keaton Stagaman, Josh Gottesman, David A. Hinds, Joyce Y. Tung, 23andMe Research Team, Stella Aslibekyan, Lucy Norcliffe-Kaufmann

## Abstract

**Importance:** The lack of information on progression, phenoconversion, and risk of dementia in a large genotyped sample impedes reliable enrichment for early interventional trials in Parkinson’s disease (PD).

**Objective:** To investigate PD penetrance, risk, motor/non-motor phenotypes, and *APOE* allele effects in *LRRK2* G2019S and *GBA* N370S carriers.

**Design:** Observational longitudinal case-control self-report survey study.

**Setting:** A US population-based study cohort enrolled in the 23andMe, Inc. and Fox Insight Genetic Substudy (FIGS) databases.

**Participants:** The total cohort included 7,586,842 participants (*n*=35,163 PD; 27% of PD cases from FIGS); 8,791 *LRRK2* G2019S carriers (565 with PD), 37,427 *GBA* N370S carriers (524 with PD), 244 dual carriers (37 with PD), and 7.5 million non-carriers (34,037 with PD).

**Exposure(s):** *LRRK2* G2019S, *GBA* N370S, *APOE* E2/E3/E4 alleles and PD polygenic risk scores (PRS).

**Main Outcome(s) and Measure(s):** Cumulative incidence of PD was estimated using Kaplan-Meier and accelerated failure time models. Relative odds of developing motor and non-motor symptoms were calculated using logistic regression models according to genetic exposure. Impact of the *APOE* alleles was estimated in a dose-dependent analysis.

**Results:** By the age of 80 years, the cumulative incidence of PD was 43% for dual carriers, 32% for *LRRK2* G2019S carriers, 6% for *GBA* N370S carriers, and 3% for non-carriers. Higher PRS was associated with increased penetrance of the variants and earlier time to PD diagnosis. Motor symptoms were similar in *LRRK2* G2019S, *GBA* N370S, and non-carriers with PD. *GBA* N370S PD was associated with the highest burden of non-motor symptoms, including REM sleep behavior disorder and cognitive/memory deficits, and *LRRK2* G2019S the lowest. *APOE* E4 dosage was associated with greater odds of developing hallucinations and cognitive decline in addition to carrier status.

**Conclusions and Relevance:** Our findings support the use of genetic screening—including *LRRK2* G2019S, *GBA* N370S, *APOE* E4, and PRS—to enrich candidate selection for neuroprotective trials and better define outcome measures based on genetic risk factors.

---

The progression of Parkinson disease (PD) is currently untreatable. For inherited monogenic forms of PD, it is possible to intervene before pathological changes become manifest clinical symptoms at a time when disease modifying therapies would be expected to have more impact.^1,2^

The most common monogenic variants associated with PD risk are located within the *LRRK2* (2% of PD cases) and *GBA* (10%) genes.^3–7^ *GBA* encodes for glucocerebrosidase (GCase), a lysosomal enzyme. GCase activity is reduced in the presence of the N370S missense variant. *LRRK2* encodes for a kinase and GTPase complex, and the common G2019S missense variant results in a gain of function in the kinase domain. Despite the growing interest in recruiting at-risk genetic cohorts, several hurdles remain to design a PD prevention study. Neither the N370S or G2019S variants are fully penetrant (i.e., not all carriers will develop PD), PD is associated with a long preclinical phase, and it is not known which key features can be used to predict progression from mild symptoms to manifest disease in monogenic forms.^8^ Without the ability to select an enriched subgroup of carriers likely to phenoconvert there is no way to shorten the period of assessment or reduce the number of participants required in an early-intervention clinical trial.

Genetic predisposition appears to be important in the pathology of PD. At least one-third of *LRRK2* carriers may have a neurodegenerative process that is independent of α-synuclein aggregation in the substantia nigra, which is the pathologic hallmark of PD^9^ Hundreds of additional genetic regions are linked to PD susceptibility.^10–12^ Polygenic risk scores (PRS) are driven by the count and weight of risk allele frequencies, and individuals at the top PRS quantiles are at particularly high risk of developing PD at an early age.^13,14^

The challenge remains to improve the precision of endpoints and to determine when to select motor versus cognitive decline as the primary outcome of interest. Although neurons in the substantia nigra are particularly susceptible to α-synuclein-mediated neurodegeneration, other regions of the brainstem, cortex, and periphery are also impacted. *GBA* PD carries more risk of dementia,^15^ whereas *LRRK2* PD appears to have a more restricted pattern of neurodegeneration and reduced risk of dementia.^14,16^ The link between the apolipoprotein E (*APOE*) E4 allele and cortical neurodegeneration is well known in Alzheimer’s dementia^17,18^ and the pathological overlap between PD and Alzheimer’s is well established, with tau and α-synuclein deposition often found in combination.^19^ More recently, it has emerged that *APOE* E4 positive carrier status may also be linked to dementia with Lewy bodies (DLB)^20^ and PD dementia;^21^ both show α-synuclein-containing Lewy bodies in the cortex.^22,23^ Whether *APOE* E4 carrier status poses a similar risk of dementia in monogenic PD is unknown.

We performed a population-based (USA) study in a cohort of 35,163 PD cases and 7,506,343 non-PD cases to investigate genotype/phenotype relationships in *LRRK2* G2019S and *GBA* N370S carriers. Our study had three main goals: (1) Define PD penetrance of the monogenic variants and whether PRS impacts PD penetrance; (2) Explore phenotypic differences among monogenic carriers; and (3) Evaluate the role of the *APOE* E4 allele in the development of dementia/hallucinations.

## Methods

### Participants

Participants with and without PD were pooled from two online prospective cohorts: 1) the 23andMe Research Database and 2) The Michael J. Fox Foundation’s Fox Insight Genetic Substudy (FIGS).^24^ Fox Insight participants were sent 23andMe genotyping kits if they reported a PD diagnosis at baseline. The data analyzed comprised June and July 2023 data cuts for 23andMe and FIGS data (https://doi.org/10.25549/bxya-6133), respectively. Both protocols were IRB approved. All participants were US-based, between 18–100 years old, and gave informed consent to participate. See eMethods for details.

### Phenotypic Data

23andMe participants who self-reported a diagnosis of PD filled a 196-item survey designed to capture past and current symptoms. Surveys within FIGS were administered upon enrollment and at predefined intervals.^24–26^ Additional longitudinal data were obtained from health and aging update surveys that followed 23andMe research participants with and without PD.^14^ Age at PD onset was defined as the minimum reported age of PD diagnosis, and discarded if the age range varied by >6 years between surveys or was <40 years. Phenotypic data were summarized by the proportion of participants that ever reported a risk factor or symptom across all timepoints and between both databases. eTable 1 lists all measures used across the 23andMe and FIGS datasets (see eMethods).

### Risk Factors and Exposures

Risk factors for PD examined included low lifetime caffeine consumption, non-smoking, family history of PD, prior traumatic brain injury (TBI), and occupational pesticide exposure. Sex (0=female, 1=male) and ancestry were derived from genotyping data.^27,28^ Level of education was binarized into less than (0) and greater than or equal to Associate degree (1).

### Genetic Data

DNA extraction and genotyping were performed on saliva samples (see eMethods). *LRRK2* G2019S and *GBA* N370S carrier status was established by the presence of pathogenic alleles determined via genotyping. Two SNPs (rs429358 and rs7412) were used to determine the *APOE* haplotype and E4 dosage. A polygenic risk score (PRS) was calculated for each participant using the allelic weights from Nalls et al.^11^ PRS including 1,805 variants. We excluded variants present in the *LRRK2* (±10 Mb window) and *GBA* (±1 Mb) loci from the PRS calculation. Principal components (PCs) of genetic ancestry for all participants and European-specific participants were computed from genotyping data (see eMethods).

### Data Analysis

Participants <40 years of age were excluded from analyses except for survival modeling; however, this did not disproportionately impact *GBA* N370S carriers, χ^2^(2) = 1.12, *p*=0.57. PD cases were stratified according to carrier status (i.e., *LRRK2* G2019S, *GBA* N370S, dual carriers, and non-carriers). Unless otherwise stated, dual carriers were excluded due to low sample sizes and *n*<5 were marked for k-anonymity.^29^ See eTable 2 for information on the statistical models, covariates, and sample sizes.

#### Penetrance estimation

Age of PD diagnosis served as our time-to-event (i.e., PD-free survival). Participants without a PD diagnosis were right censored at the age of their most recent survey completion. To define PD penetrance incident to genetic exposure, we analyzed participants with pre-existing diagnosis of PD.^14,30,31^ We used the Kaplan-Meier method to estimate PD-free survival probability as a function of carrier status across four levels: 1) *LRRK2* G2019S, 2) *GBA* N370S, 3) dual carriers, and 4) non-carriers. Because the effect of carrier status violated the proportional hazard assumption of Cox regression based on Schoenfeld residuals and time, χ^2^(3) = 43.25, *p*=2.18 × 10^−09^, we used Weibull accelerated failure time (AFT) models to explore the effect of PRS and carrier status on PD-free survival probability. We performed AFT modeling with and without the covariates of sex and the first 10 ancestry PCs. We repeated the AFT models excluding non-Europeans using sex and the first five European ancestry PCs as covariates. Predicted survival probabilities and cumulative incidence were generated for males with sample means for PRS and ancestry PCs. Male sex was arbitrarily chosen for predicted estimates; however, we also report female cumulative incidence of PD in the eMethods.

#### PD Risk

The risk of developing PD was examined using two logistic regression analyses.^14,32^ First, we modeled the relative odds of developing PD as the interaction of carrier status, which included dual carriers, and PRS group: low PRS (1-25%), middle PRS (25-75%), and high PRS (75-100%). Non-carriers with middle PRS served as the reference group. Second, we modeled the relative odds of developing PD as the interaction of carrier status and PRS decile; non-carriers with median PRS (5^th^ decile) served as the reference group. Normalized age (*M*=0, *SD*=1), sex, and the first 10 ancestry PCs were used as covariates. Analyses were performed including and excluding non-Europeans (in which case the first five European ancestry specific PCs were used as covariates). We estimated deviations from additivity using ANOVA. Predicted odds ratios were generated for males with sample means for age and ancestry PCs.

#### Symptomatic burden

Logistic regression models estimated the relative odds of reporting each symptom as a function of carrier status. Sufficient sample sizes were unavailable for most symptoms in dual carriers; therefore, the dual carrier models are presented in the eMethods. Non-carriers served as the reference group, and sex and education were used as covariates. Using previously described methods,^14^ we used the prevalence of symptoms to visually reconstruct patterns of neurodegeneration in five brain regions including the substantia nigra (motor), pons (RBD), medulla/lower brainstem (autonomic), cerebral cortex and limbic areas (cognitive/memory/psychotic), and olfactory bulb (hyposmia; see eMethods). We adjusted the *p*-values using false discovery rate and used post-hoc Tukey-corrected pairwise comparisons to examine differences between carrier status.

#### APOE status and cognitive/memory/psychotic symptoms

The impact of *APOE* haplotype and E4 dosage was assessed across four cognitive/memory/psychotic symptoms: concentration, hallucinations, general memory, and memory for dates. Logistic regressions were used to model the relative odds of each symptom as a function of carrier status and *APOE* haplotype (E3/E3 as reference group). Logistic regression was used to model the relative odds of each symptom as a function of *APOE* E4 dosage (0, 1, 2) as a continuous variable, carrier status (non-carriers as reference group), and their interaction. Sex and education served as covariates.

#### Software

All analyses were performed using R (v. 4.1.2).^33^ Survival analyses were performed using R packages *survival*^34^ and *flexsurv*.^35^ Figures were prepared using R packages *ggplot2*^36^ and *patchwork*.^37^

## Results

### PD Penetrance was Greatest in Dual Carriers

The cohort demographics and symptoms are summarized in Table 1 (see eTable 3 for total sample sizes), and the study flow in Figure 1. The mean age of PD diagnosis for dual carriers was 60.1 years (*SEM*=1.6 years), 61.4 (*SEM*=0.5) for *LRRK2* G2019S, 59.1 (*SEM*=0.5) for *GBA* N370S, and 61.5 (SEM=0.1) for non-carriers. The Kaplan-Meier analysis, which included *n*=32,143 PD cases and *n*=7,343,120 non-PD controls, is shown in eFigure 1. PD-free survival was different between carrier groups (log-rank test χ^2^(3) = 6,316.00, *p*<.001). The cumulative incidence of PD at age 80 was highest in dual carriers (43%), followed by *LRRK2* G2019S (32%), *GBA* N370S (6%), and non-carriers (3%; see eTable 4). The Weibull AFT models without covariates showed similar results (see eTable 5 and eFigure 2).

**Table 1.**
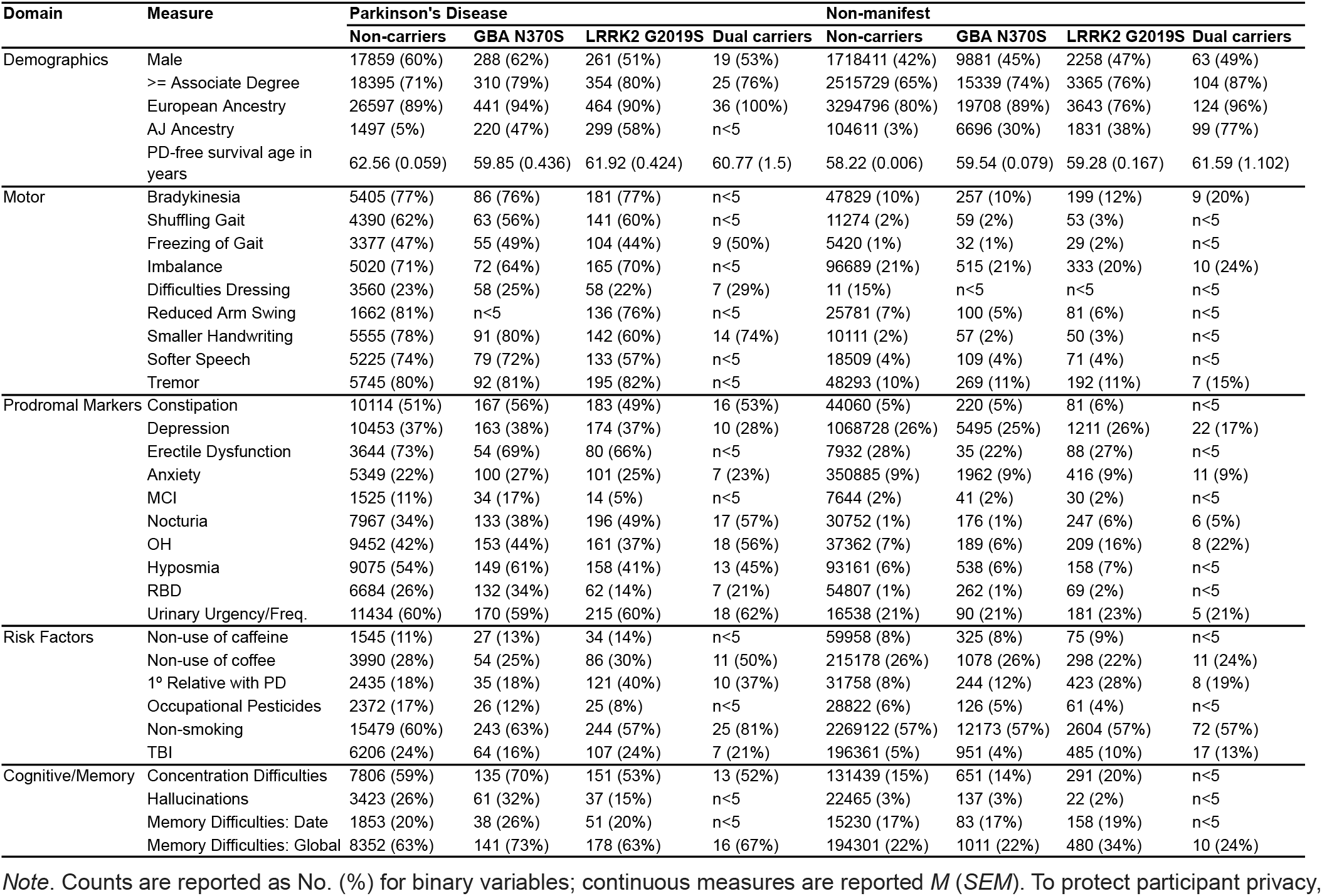
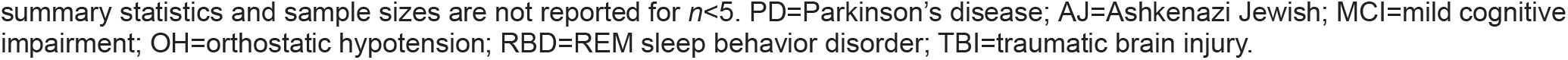
Demographics and self-reported symptoms and risk factors in participants ≥ 40 years of age.

**Figure 1.**
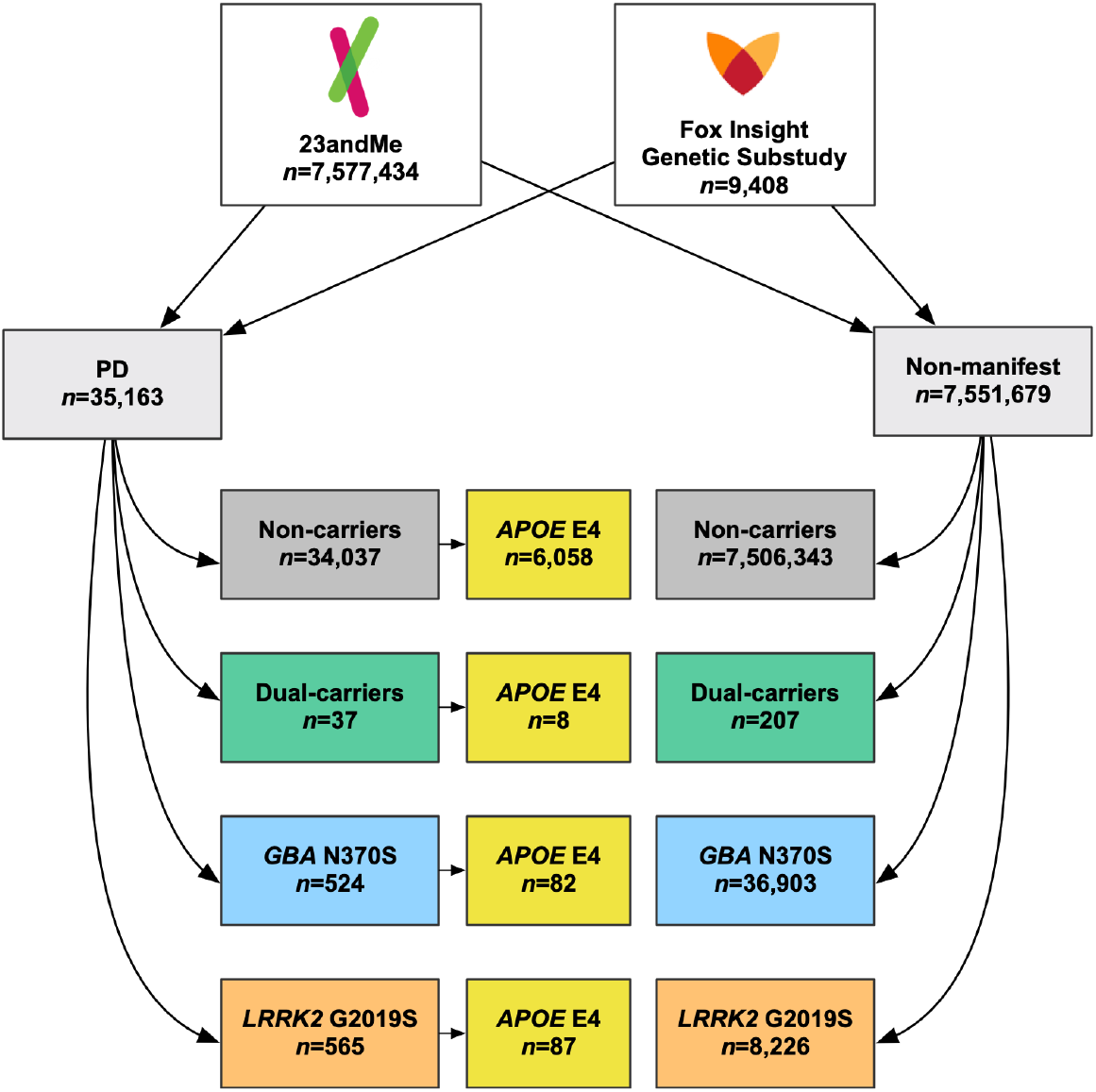
Participant flow diagram. Participants from the 23andMe Research Cohort and the Fox Insight Genetic Substudy (July 2023 data cut) with a clearly defined PD status and available genotyping data were combined into a mega-analysis of PD symptoms, PD-free survival, PD risk, and PRS. Sample sizes in the colored boxes represent total possible sample sizes used for the analyses presented. *LRRK2* G2019S, *GBA* N370S, dual carriers, and *APOE* E4 carrier groups were defined as carrying at least one pathogenic variant.

### High PRS Increases Risk of PD

There was a significant effect of PRS on PD-free survival probability (Figure 2). A greater PRS accelerated the time to PD diagnosis across all carrier groups (*TR*=.921 [.920 .923], *p*<.001; see eTable S5). An increase in 1 *SD* of PRS predicted an earlier age at 50% survival probability by 6.54 [.52 12.56] years for dual carriers, 7.00 [5.26 8.73] years for *LRRK2* G2019S, 8.97 [6.66 11.23] years for *GBA* N370S, and 10.23 [9.35 11.11] years for non-carriers. Relative to non-carriers with middle PRS, having a high PRS increased the odds of developing PD 45-fold for dual carriers (OR=45.73 [26.01 80.38]), 22-fold for *LRRK2* G2019S carriers (*OR*=22.13 [19.05 25.71]), 5-fold for *GBA* N370S carriers (*OR*=4.92 [4.31 5.62]), and doubled for non-carriers (*OR*=2.01 [1.96 2.06]; see Figure 2C and eTable 6). The odds of developing PD were highest for *LRRK2* and dual carriers regardless of PRS. The risk of PD overlapped in non-carriers with high PRS and *GBA* N370S carriers with low PRS. A similar impact of high PRS on disease risk was observed in the decile analysis (see Figure 2D; eTable 7). Departure from additivity was observed in the PRS group (low, middle, high) model (ΔDeviance(6)=14.47, *p*=.02 via χ2 test) but not in the decile model (ΔDeviance(18)=23.05, *p*=.19). We observed similar results when excluding those with non-European ancestry (see eTables 7–11 and eFigures 2–4).

**Figure 2.**
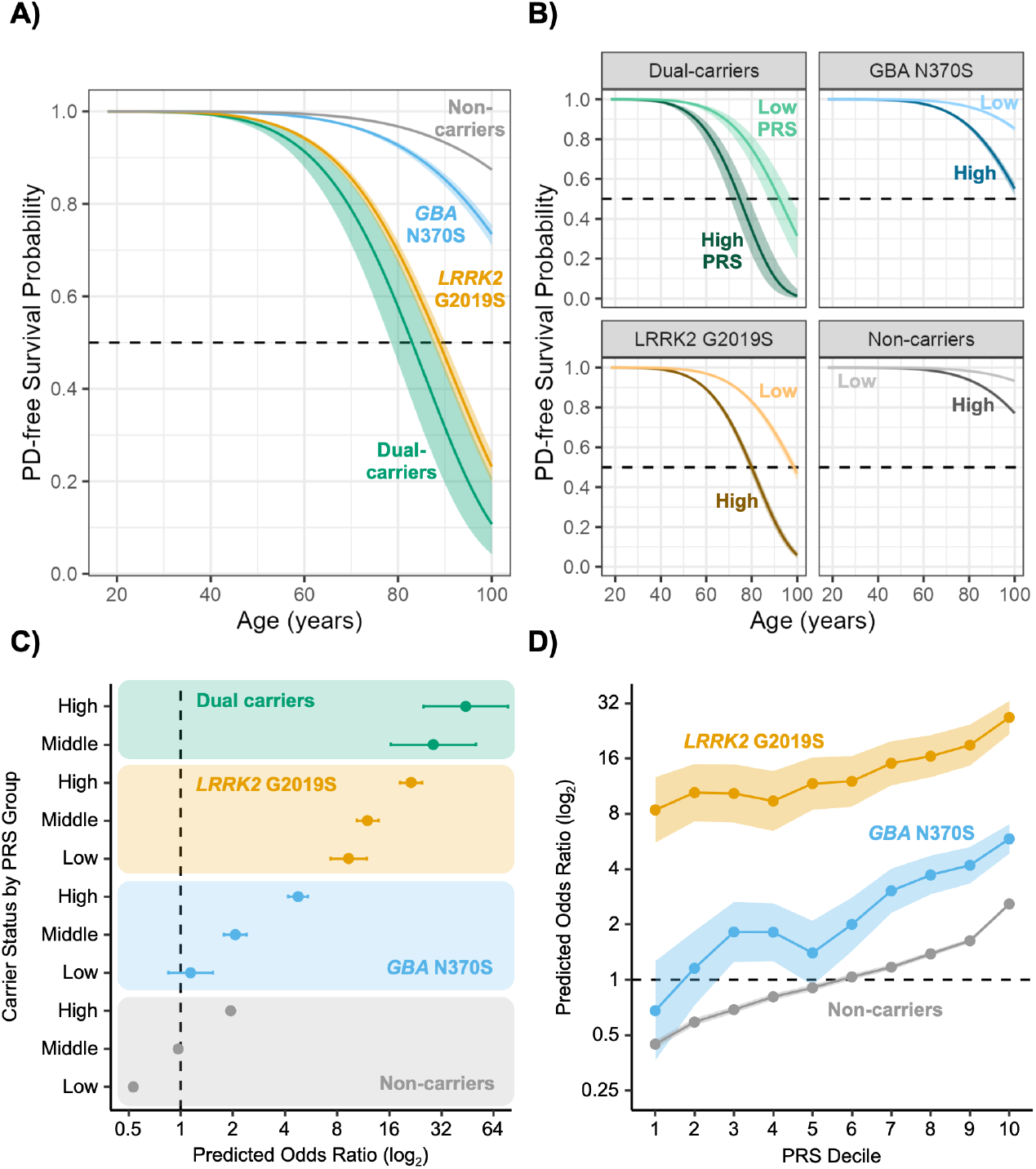
Dual *LRRK2* G2019S/*GBA* N370S carriers have the greatest PD penetrance and risk of developing PD. **A)** Curves represent predicted PD-free survival probability for males using the sample means for PRS and PCs. **B)** Curves represent predicted PD-free survival probability for low (10%) and high (90%) PRS, males, and mean PCs. Shading depicts 95% CI. **C)** Forest plot of predicted odds ratios indicating a positive association between PD risk and PRS within carrier groups. PRS quantiles defined groups: low (bottom 25%), middle (between 25-75%), and high (upper 25%). Dual carriers with low PRS were removed from the plot due to small sample size. Non-carriers with middle PRS was the reference group. Predicted odds ratios were computed for males using sample means of ancestry PCs and age. **D)** The predicted relative odds of developing PD according to PRS decile. Predicted odds ratios were computed as described in C, but non-carriers at the fifth decile were the reference group. All error bars and shading reflect 95% CIs.

### In PD Participants, Greater Prevalence of Prodromal Symptoms, Cognitive Impairment, and Hallucinations in *GBA* N370S relative to *LRRK2* G2019S Carriers

The prevalence of symptoms in participants with PD within each neuroanatomical domain for *LRRK2* G2019S, *GBA* N370S, and non-carriers are shown in Figure 3 (dual carrier results reported in eMethods). Odds ratios are presented in eTable 12 and post hoc pairwise Tukey tests in eTable 13. The prevalence of PD motor symptoms was similar across all carrier groups, with the exception that fewer *LRRK2* G0219S PD reported hypophonia (softer speech) and micrographia (smaller handwriting) than *GBA* N370S PD and non-carriers with PD.

**Figure 3.**
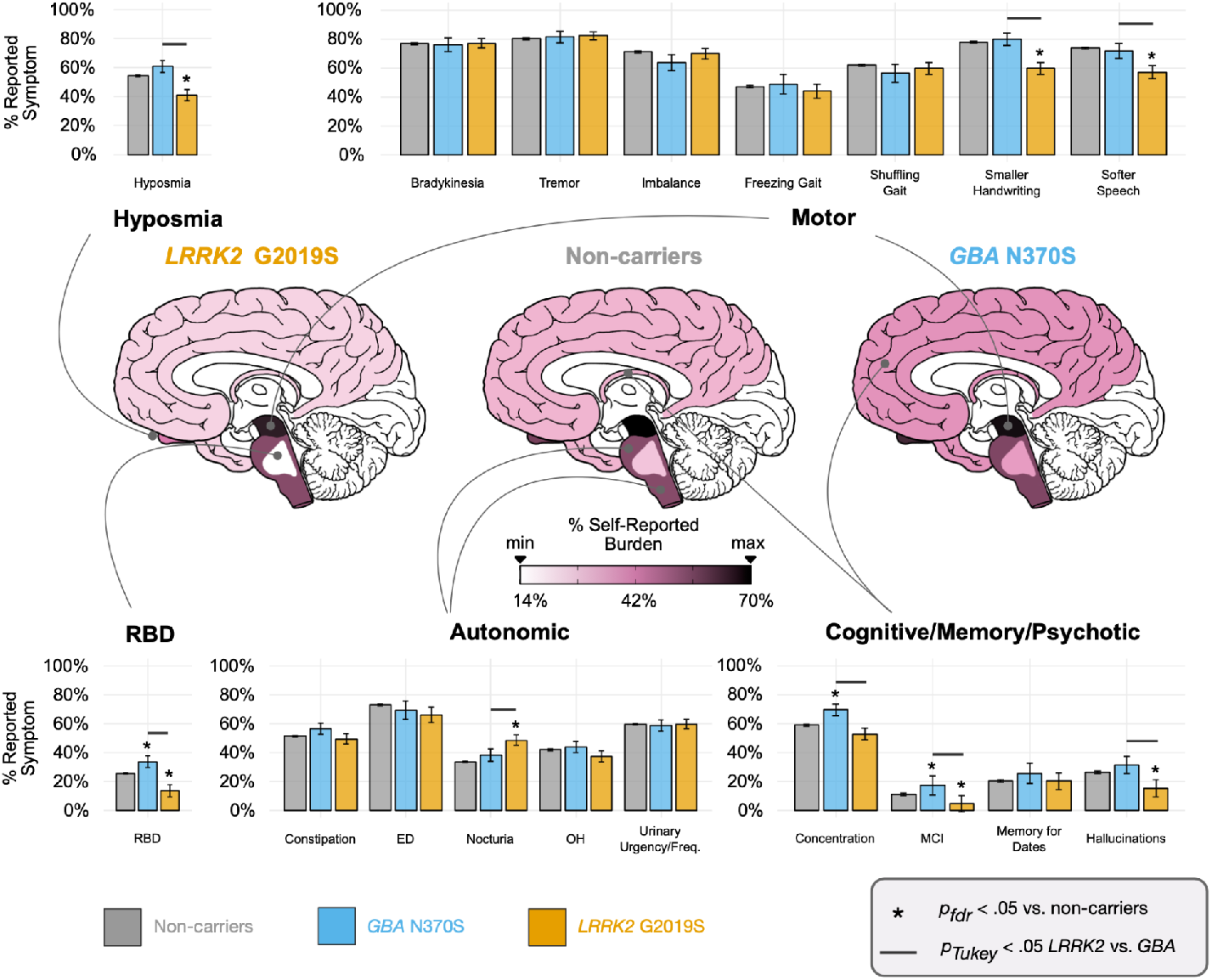
Self-reported symptom prevalence in *LRRK2* G2019S carriers with PD, *GBA* N370S carriers with PD, and non-carriers with PD (idiopathic PD). Symptoms across questionnaires were aggregated into six domains and brain regions were shaded in approximated neuroanatomical regions according to average reported symptom burden across symptoms within domain: motor (substantia nigra), autonomic (brain stem excluding regions of the pons), cognitive/memory/psychotic (cerebral cortex and limbic areas), hyposmia (olfactory bulb), REM sleep behavior disorder (RBD; areas of the pons). The false discovery rate (FDR) was adjusted in carrier group comparisons to non-carriers within symptom domains. Comparisons between *LRRK2* G2019S and *GBA* N370S carriers were adjusted with Tukey’s honestly significant difference tests. Dual carrier results are presented in the eMethods. Error bars are *SE*. Descriptive statistics are not reported for measures with *n*<5 due to 23andMe data privacy policies.

The prevalence of non-motor autonomic symptoms was similar between carrier groups, with the exception that *LRRK2* G2019S PD reported more nocturia than *GBA* N370S PD and non-carriers with PD. Compared to idiopathic (non-carrier) PD, the prevalence of RBD and hyposmia were higher in *GBA* N370S PD, and lower in *LRRK2* G2019S PD. Cognitive symptoms were different among genetic subtypes. *GBA* N370S PD had the highest prevalence of concentration and memory problems, were more likely to be diagnosed with MCI, and were more likely to report hallucinations. *LRRK2* G2019S PD reported significantly fewer cognitive symptoms and less hallucinations. Cognitive symptoms were not more severe in dual carriers (see eTables 14–15 and eFigure 5).

### *APOE* E4 Dosage Increases Risk of Cognitive and Psychotic Symptoms in *LRRK2* G2019S and *GBA* N370S Carriers with PD

The presence of the *APOE* E4 allele status was associated with higher prevalence of hallucinations (see eTable 16 and Figure 4A). Each additional copy of the *APOE* E4 allele conferred a 16% increase in the odds of reporting hallucinations (see eTable 17 and Figure 4B). Similarly, *APOE* E4 dosage was associated with greater odds of developing memory and concentration issues after adjusting for sex and education. We did not observe any interactions between *APOE* E4 dosage and carrier status, suggesting that *APOE* E4 dosage contributes an additive risk (on the log odds scale) to cognitive/memory/psychotic symptoms in *LRRK2* G2019S and *GBA* N370S carriers with PD; however, we were underpowered to fully test this hypothesis (see eTable 18 for sample sizes).

**Figure 4.**
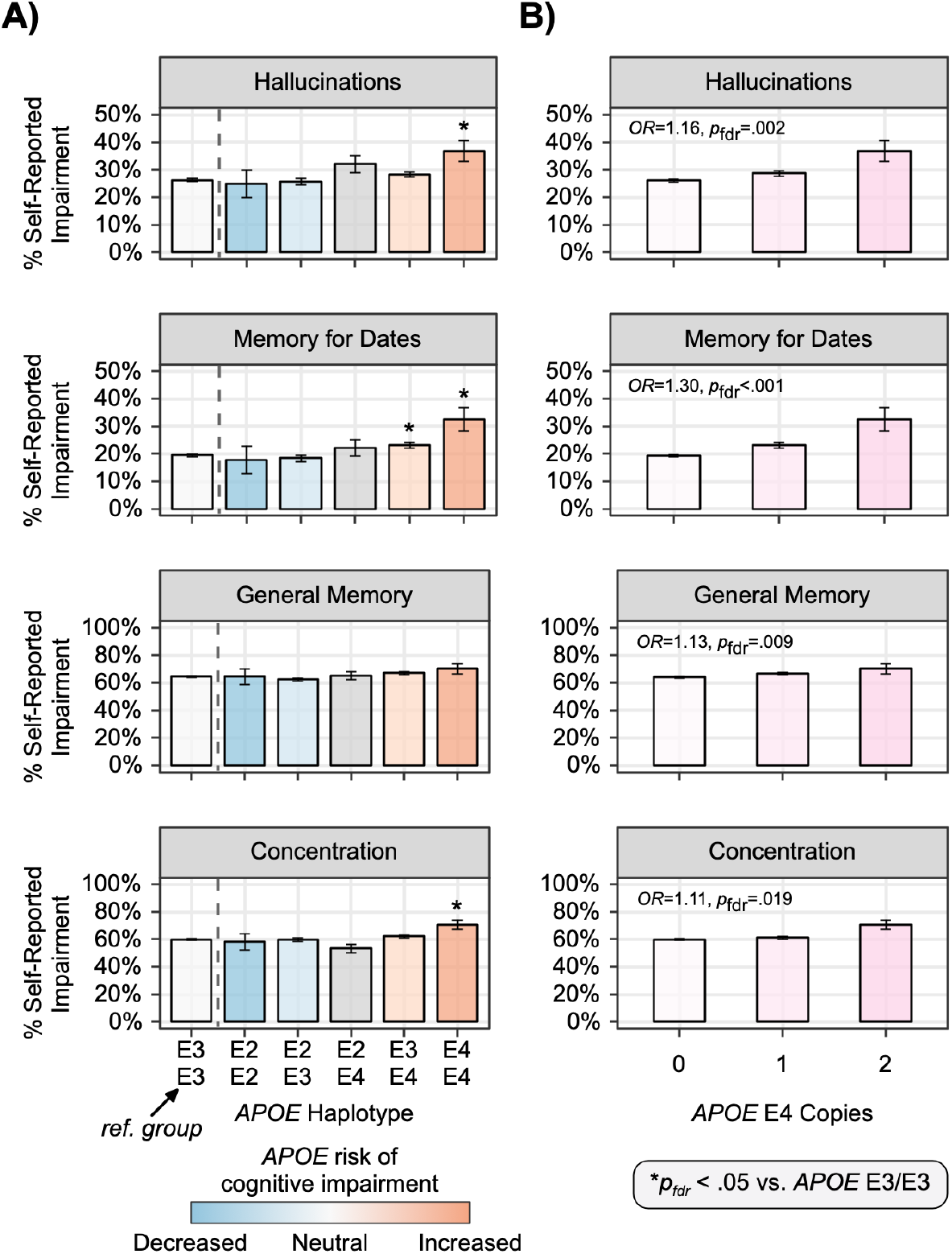
The effect of *APOE* haplotype and E4 dosage on cognitive/memory/psychotic symptoms in individuals with PD. A) The percentage of participants with PD with cognitive/memory/psychotic symptoms across possible *APOE* haplotypes collapsed across carrier status. Stars indicate significantly greater relative odds for reporting symptoms compared to *APOE* E3/E3 individuals (i.e., reference group). Bars are color coded by hypothesized risk of cognitive impairment due to *APOE* haplotype; E2/E4 carriers carry one protective (E2) and one risk variant (E4) and are therefore colored grey. B) Bar graphs show a positive association between *APOE* E4 dosage and percentage of cognitive/memory/psychotic symptoms in participants with PD collapsed across carrier status. Error bars are *SE*.

## Discussion

This population-based analysis demonstrates unequivocally that *LRRK2* G2019S is five-times more penetrant than the *GBA* N370S variant, but the penetrance of PD is highest for dual carriers. PRS modifies PD risk, such that *GBA* N370S carriers with low PRS have similar risk of developing PD as non-carriers with median PRS. Carriage of *APOE* E4 variants increases the risk of developing cognitive impairment in PD, whereas the *LRRK2* G2019S variant reduces the risk of hallucinations and dementia.

We show that by the age of 80 years, 43% of dual carriers, 32% for *LRRK2* G2019S carriers and 6% of *GBA* N370S carriers developed PD, compared to 3% of non-carriers. Although dual carriers have a high cumulative incidence of PD, the combination of both variants is extremely rare, and accounts for only 1:33,000 PD cases. *LRRK2* G2019S, although more highly penetrant, has a carrier rate of 1:1,100 PD cases, whereas *GBA* N370S is less penetrant but has a more frequent carrier rate 1:200. Although these single variants are common in monogenic PD,^6^ additional risk markers of imminent phenoconversion are needed to enrich early-interventional clinical trials. *GBA* variants have also been linked to cases of dementia with Lewy bodies and multiple system atrophy,^38^ which underlies the importance of finding biomarkers to optimize the distinction between the different α-synucleinopathies in the early stages.

Our findings show the risk of developing cognitive impairment is highest in *GBA* N370S carriers. Based on the anatomical models, we show that while *GBA* N370S is considered a “mild” variant, it is associated with widespread neuronal loss beyond the substantia nigra. This is consistent with clinical cohorts^39–45^ and pathology studies showing Lewy body pathology throughout the cortex in *GBA* carriers.^22,46–48^ A novel finding in our study was that each additional copy of *APOE* E4 conferred between 11-30% increase in the odds of developing hallucinations and cognitive impairment in PD. This adds to the risk of a more severe cognitive phenotype in *GBA* N370S carriers, and may be useful when tracking progression.^22,49^ In contrast to others,^49^ we did not find evidence suggesting that the *APOE* E2 allele was protective.

Our data add to the growing evidence that *LRRK2* appears to reduce the risk of non-motor symptoms. First, we show that *LRRK2* G2019S PD has the lowest prevalence of hallucinations, RBD and hyposmia. Second, we show that dual carriers do not report more severe symptoms than *GBA* carriers across all non-motor symptom domains (see eFigure 5). This aligns with other reports that suggest the presence of the gain of function *LRRK2* G2010S variant offsets the down regulation of GCase.^50–52^ While α-synuclein Lewy body cortical involvement may not always predict dementia,^53^ it is worthwhile noting that at least one-third of *LRRK2* carriers do not have evidence of α-synuclein seeding in the CSF^9^ and do not show CNS Lewy bodies at autopsy^54,55^ which suggests a restricted neurodegenerative process that is independent of α-synuclein and slower progressing.^14,56–59^

A major obstacle for early-interventional disease-modifying clinical trials is lack of enrichment.^60^ One promising enrichment strategy is PRS. PRS can heighten risk of phenoconversion at an early age,^11,61,62^ including in monogenic cases,^13,14,63^ and aid in balancing treatment arms^64^. As shown by our data, PRS affects PD penetrance more than sex, low caffeine intake, non-smoking, and TBI.^14^ *GBA* N370S carriers at the top decile for PRS experienced a six-fold increase in the relative odds of developing PD compared to non-carriers with median PRS, which may help reduce the number of participants required for *GBA* trials. An even greater risk was seen in *LRRK2* G2019S carriers, who experienced nearly 30-fold increase at the top PRS decile relative to non-carriers with median PRS. However, given the rarity of the *LRRK2* G2019S variant, too strict a PRS-based enrichment for *LRRK2* trials would make it difficult to recruit a sufficient number of candidates in a timely manner.

There are some limitations to our study. The 23andMe cohort is unique, assembled by virtue of study participants being interested in health-related genetics, and likely more affluent. The PD status and symptom prevalence in both the Fox Insight and 23andMe databases is based on self-report, but appears to have excellent correlation with clinical evaluations.^65–67^ To create a more homogenized cohort, we restricted the age of PD diagnosis to ≥40 years that eliminated a small minority of early onset cases. We observed a lower predicted cumulative incidence of PD than previously reported.^14,68–70^ This lower PD incidence may be the result of 1) using a large population-based non-manifest cohort and 2) adjusting for genetic ancestry PCs and PRS that previous studies did not perform.

## Conclusion

Our findings support the use of genetic screening and PRS to enrich candidate selection for neuroprotective trials. The distinct pathways affected by *LRRK2* and *GBA* mutations may offer insights into the pathobiology of PD and strategies for treating progression.

## Supporting information

eTable

eMethods

## Data Availability

Model outputs for all logistic regressions and survival models are provided as eTables. Individual-level data from 23andMe are not publicly available due to participant confidentiality and in accordance with the IRB-approved protocol under which the study was conducted. The Fox Insight Genetic Substudy participant data are available through Fox DEN (https://foxden.michaeljfox.org). No custom code or software was generated as part of the study. Details of all software packages used for data processing and analysis may be found in the "Methods" section.

## Acknowledgements

We would like to thank Mina Kmiecik for data illustrations, Madeleine Wetzel and Helen M. Rowbotham, and the research participants and employees of 23andMe for making this work possible. The following members of the 23andMe Research Team contributed to this study: Adam Auton, Elizabeth Babalola, Robert K. Bell, Jessica Bielenberg, Ninad S. Chaudhary, Zayn Cochinwala, Sayantan Das, Emily DelloRusso, Payam Dibaeinia, Sarah L. Elson, Nicholas Eriksson, Chris Eijsbouts, Teresa Filshtein, Davide Foletti, Will Freyman, Zach Fuller, Julie M. Granka, Chris German, Éadaoin Harney, Alejandro Hernandez, Barry Hicks, M. Reza Jabalameli, Ethan M. Jewett, Yunxuan Jiang, Sotiris Karagounis, Katelyn Kukar, Alan Kwong, Keng-Han Lin, Yanyu Liang, Bianca A. Llamas, Aly Khan, Steven J. Micheletti, Matthew H. McIntyre, Meghan E. Moreno, Priyanka Nandakumar, Dominique T. Nguyen, Jared O’Connell, Steve Pitts, G. David Poznik, Alexandra Reynoso, Shubham Saini, Morgan Schumacher, Leah Selcer, Anjali J. Shastri, Suyash Shringarpure, Teague Sterling, Qiaojuan Jane Su, Vinh Tran, Xin Wang, Wei Wang, Catherine H. Weldon, Amy L. Williams, Peter Wilton.

