## Supplementary material for "Monogenic Parkinson’s Disease and the Impact of *APOE* E4: A Case-Control Study": eMethods

### Participants

Study recruitment occurred by voluntary participation, website advertisement, and targeted email. 23andMe research participants who self-reported a diagnosis of Parkinson disease (PD) or had reviewed their PD Genetic Health Risk report (i.e., were aware of their *LRRK2* G2019S and *GBA* N370S carrier status) and agreed to being recontacted for research were recruited into the Fox Insight Genetic Substudy (FIGS) study that ran from 2017–2021 (Gottesman et al., 2024). 23andMe genotyping and FIGS survey responses were de-identified and linked through an encrypted key.

23andMe research participants provided informed consent and volunteered to participate in the research online, under a protocol approved by Ethical & Independent (E&I) Review Services–an external IRB that is accredited by the Association for Accreditation of Human Research Protection Programs. As of 2022, E&I Review Services is part of Salus IRB (https://www.versiticlinicaltrials.org/salusirb).

FIGS participants were 18 years or older and provided informed consent via the Fox Insight website (WCG IRB IRB#: 120160179, Legacy IRB#: 14–236, Sponsor Protocol Number: 1, Study Title: Fox Insight) (Gottesman et al., 2024). FIGS data used in the preparation of this article were obtained from the Fox Insight database https://foxden.michaeljfox.org using the July 2023 data cut. For up-to-date information on the study, visit https://foxden.michaeljfox.org.

### Phenotypic Data

We first ran a qualitative analysis to determine the overlapping questions from surveys within the 23andMe database and Fox Insight, and harmonized data by matching survey responses (see eTable 1). Categorical variables were converted into binary measures (yes/no). Self-reported motor symptoms assessed included: tremor, bradykinesia, shuffling gait, freezing gait, imbalance/falls, reduced arm swing, micrographia (small handwriting), and hypophonia (softer speech). Non-motor symptoms included constipation (<3 bowel movements/week), nocturia (>two times/night), neurogenic orthostatic hypotension, hyposmia (very poor/poor sense of smell), increased urinary frequency/urgency, erectile dysfunction (males only), diagnosed or suspected REM sleep behavior disorder (RBD), anxiety, and depression. Cognitive symptoms included poor concentration (i.e., difficulty staying focused/paying attention), trouble remembering the date/day/month, and generalized memory problems. Participants were also asked whether they experienced hallucinations or had been diagnosed with mild cognitive impairment.

### Genotyping

DNA extraction and genotyping were performed on saliva samples by Clinical Laboratory Improvement Amendments-certified and College of American Pathologists-accredited clinical laboratories of Laboratory Corporation of America. Samples were genotyped on one of five genotyping platforms. The V1 and V2 platforms were variants of the Illumina HumanHap550+ BeadChip and contained a total of approximately 560,000 SNPs, including about 25,000 custom SNPs selected by 23andMe. The V3 platform was based on the Illumina OmniExpress + BeadChip and contained a total of about 950,000 SNPs and custom content to improve the overlap with our V2 array. The V4 platform is a fully custom array and includes a lower redundancy subset of V2 and V3 SNPs with additional coverage of lower-frequency coding variation, and about 570,000 SNPs. The V5 platform is an Illumina Infinium Global Screening Array of about 640,000 SNPs supplemented with about 50,000 SNPs of custom content. From the total sample size of *n*=7,586,842 participants, 0.24% were genotyped on V2, 2.24% on V3, 15.73% on V4, and 81.79% on V5.

Greater than 99% of the genotyping results agreed with the Sanger sequencing results for *LRRK2* G2019S, *GBA* N370S, and the two SNPs (rs429358 and rs7412) used to determine the *APOE* E4, E3, and E2 haplotype. Likewise, these variants exhibited >99% reproducibility and repeatability on the genotyping platform (23andMe, Inc., 2023).

**Imputation Reference Panels**

Variants were imputed using a reference panel comprised of three other reference panels: the publicly available Human Reference Consortium (HRC) and UK BioBank (UKBB) 200K Whole Exome Sequencing (WES) reference panels, and the 23andMe reference panel. HRC data were downloaded from the European Genome-Phenome Archive at the European Bioinformatics Institute (accession EGAD00001002729). Variants were converted to hg38 and excluded if their new positions were on a different chromosome. Variants were then re-phased using SHAPEIT4 (https://odelaneau.github.io/shapeit4/). Finally, singletons were excluded. The final HRC reference panel included 27,165 samples and 39,057,040 SNPs (no indels).

The UKBB 200K WES reference panel was built following a recently published approach (Barton et al., 2021). UKBB genotyping data included 486,633 samples and 681,298 variants, and was lifted from hg19 to hg38 and re-phased using SHAPEIT4. UKBB 200K WES data included 200,643 whole exome sequenced samples (mostly British ancestry) and more than 22M variants (https://www.ukbiobank.ac.uk/media/cfulxh52/uk-biobank-exome-release-faq_v9-december-2020.pdf). Multi-allelic variants were split into bi-allelic variants using bcftools. Genotypes with GQ<20 were set to missing. Variants with >20% missingness, a minor allele count of 0, or an inbreeding coefficient < -0.3 (high heterozygosity) were removed. After QC, 17,975,023 UKBB WES variants remained. For the 199,815 samples with both WES and genotype array data, the variants were merged and re-phased using SHAPEIT4. After excluding singletons, the UKBB 200K WES reference panel included 199,815 samples and 17,082,588 variants (16,115,376 SNPs and 967,212 indels).

The 23andMe reference panel included 12,217 samples from: consented 23andMe customers, the 1000 Genomes Project (Auton et al., 2015), Syndip (Li et al., 2018), the Genotype-Tissue Expression (GTEx) project v8 (Aguet et al., 2019), the Human Genome Diversity Project (HGDP) (Bergström et al., 2020), and the Simons Genome Diversity Project (SGDP) (Mallick et al., 2016). All samples (except for those from GTEx v8) were consented for research as of 2020-01-18, had sufficient depth of coverage (greater than the median - 3*MAD (median absolute deviation) within each cohort), had contamination < 0.05 as estimated by *verifybamid*, had <0.05 chimeric reads, had median insert size >= 250 bp, had r2 with genotyping array >= 0.8, and had >= 3M SNPs called. GTEx v8 samples were sometimes slightly outside these bounds, but were included due to their value in eQTL mapping. All samples were aligned to GRCh38_full_analysis_set_plus_decoy_hla.fa9 and duplicate marked. Datasets sequenced before 01/01/2019 were re-aligned using an in-house pipeline consisting of bwa mem 0.7.15-r1140 alignment and duplicate marking with samblaster v0.1.24. Datasets sequenced after 01/01/2019 were processed using a well-known public pipeline from the Broad Institute (Aguet et al., 2019) that combined bwa mem 0.7.15-r1140 alignment, Picard MarkDuplicates

2.15.0, and BQSR with GATK 4.beta.5. Variants were called in each individual sample using DeepVariant-0.8.0 11 to produce GVCFs. The GVCFs were then joint-called using GLnexus-1.2.3 12. Singletons were removed, genotypes with GQ<20 were set to missing, variants with >20% missingness (after the GQ filter) were removed, and variants with >30% excess heterozygosity were removed. Finally, variants were phased using SHAPEIT4. SHAPEIT4 also imputed missing genotypes and produced a final panel without missingness. The final 23andMe reference panel included 12,217 samples and 82,078,539 variants (73,852,355 SNPs + 8,226,184 indels).

**Imputation**

Imputation was performed using Beagle 5 (Browning et al., 2018). Because participants were genotyped on one of the five genotyping platforms (v1 to v5), imputation was performed independently for each platform. Similarly, participants were classified into one of five ancestry-based populations as described previously (https://www.23andme.com/ancestry-composition-guide/) and imputation was performed independently for each population. We used a two-step imputation process: first combining results from the HRC and 23andMe reference panels, then combining the HRC+23andMe results with UKBB 200K WES results. In the first step, there were 85,099,656 variants found in either the HRC or 23andMe reference panels. HRC-specific variants (n=3,021,117) were imputed using all HRC reference panel samples. 23andMe-specific variants (n=46,042,616) were imputed using all 23andMe reference panel samples. Variants found in both HRC and 23andMe reference panels (hereafter referred to as HRC+23andMe, n= 36,035,923) were imputed using all HRC+23andMe reference panel samples. In the second step, there were 99,675,338 variants found in either the HRC+23andMe or UKBB 200K WES reference panels. HRC+23andMe-specific variants were imputed as described in step 1 (n=82,592,750). UKBB 200K WES-specific variants were imputed using the UKBB 200K WES reference panel (n= 14,575,682). Variants found in both the HRC+23andMe and the UKBB 200K WES reference panels (n= 2,506,906) were imputed in each panel separately and the result with the higher imputation quality was used. Imputation quality was taken as the sample size-weighted imputation R2 value across genotyping platforms and populations.

**Polygenic Risk Score (PRS) Calculation**

The 1,805 variant PRS (Nalls et al., 2019) was a weighted sum of risk allele counts for the included SNPs that were matched using CPRA (chromosome, position, reference allele, alternative allele) format and harmonized to 23andMe imputation panel (Hartwig et al., 2016). We removed SNPs with imputation $R^2<0.5$ or a difference in MAF>30% between the Nalls et al. (2019) and 23andMe variants. Of the total 1,805 SNPs, 26 were unmatched, 46 SNPs were removed due to the *LRRK2* proximity, and 16 SNPs were removed due to *GBA* proximity, resulting in a total of 1,717 variants for the final PRS used for analyses. PRS was normalized ($M$=0, $SD$=1) using the entire database of 23andMe and FIGS participants at time of database freeze (n=11,178,995).

**Principal Components of Ancestry**

Principal components (PCs) for the full cohort were derived from a principal component analysis performed on one million randomly selected participants from 23andMe’s Research Cohort using 63,528 high quality genotyped variants present across all five genotyping platforms. Loadings for participants not included in the analysis were obtained by projection, combining the eigenvectors of the analysis and the SNP weights. Similarly, European-specific PCs were derived from a principal component analysis performed on one million randomly selected participants from European ancestry, using the same set of 63,528 high quality genotyped variants. Genetic ancestry PCs, including both all ancestry and European-specific PCs, were then normalized ($M$=0, $SD$=1) using the entire database of 23andMe and FIGS participants at time of database freeze ($n$=11,178,995).

**Symptomatic Burden**

Similar to our previous model recapitulating the neurobiology of PD (Kmiecik et al., 2024), we mapped symptoms to suspected regions of neurodegeneration across the brain and shaded in approximated neuroanatomical regions according to average reported symptom burden across the following domains: substantia nigra (motor: bradykinesia, tremor, imbalance, shuffling gait, freezing gait, reduced arm swing, smaller handwriting, softer speech), brain stem excluding regions of the pons (autonomic: constipation, erectile dysfunction, nocturia, orthostatic hypotension, increased urinary urgency/frequency), cerebral cortex and limbic areas (cognitive/memory/psychotic: concentration difficulties, mild cognitive impairment diagnosis, difficulties with memory for dates, hallucinations), olfactory bulb (hyposmia), areas of the pons (RBD). We adjusted the p-values using false discovery rate within each symptom domain.

### Supplementary eTables

All supplementary tables via an Excel workbook are available for download.

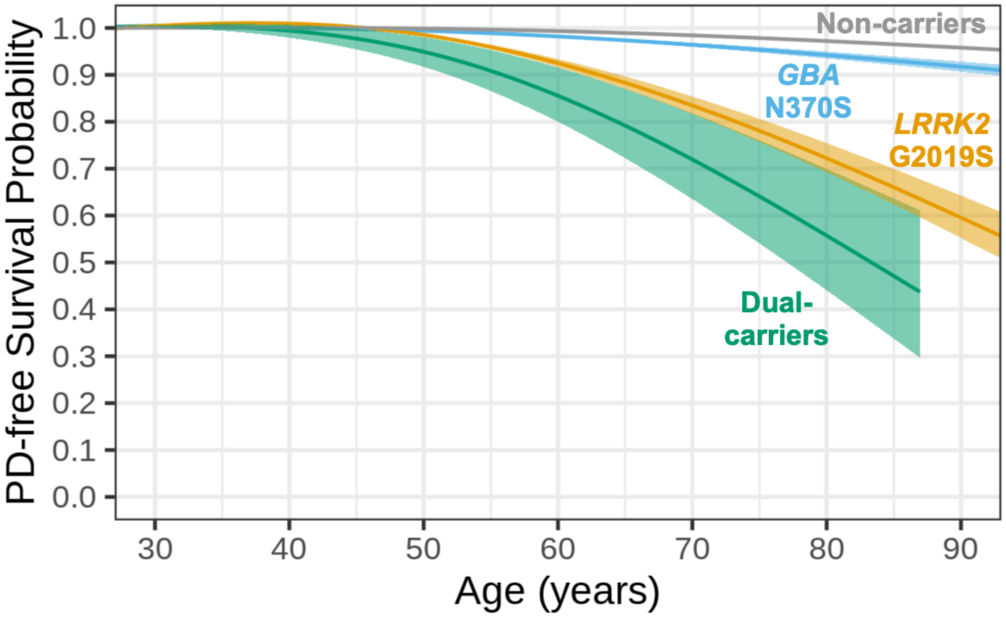


**eFigure 1. Kaplan-Meier estimated PD-free survival probabilities stratified by carrier status across all ancestries.** Curves were smoothed using generalized additive modeling to protect participant privacy. Shading denotes 95% confidence intervals.

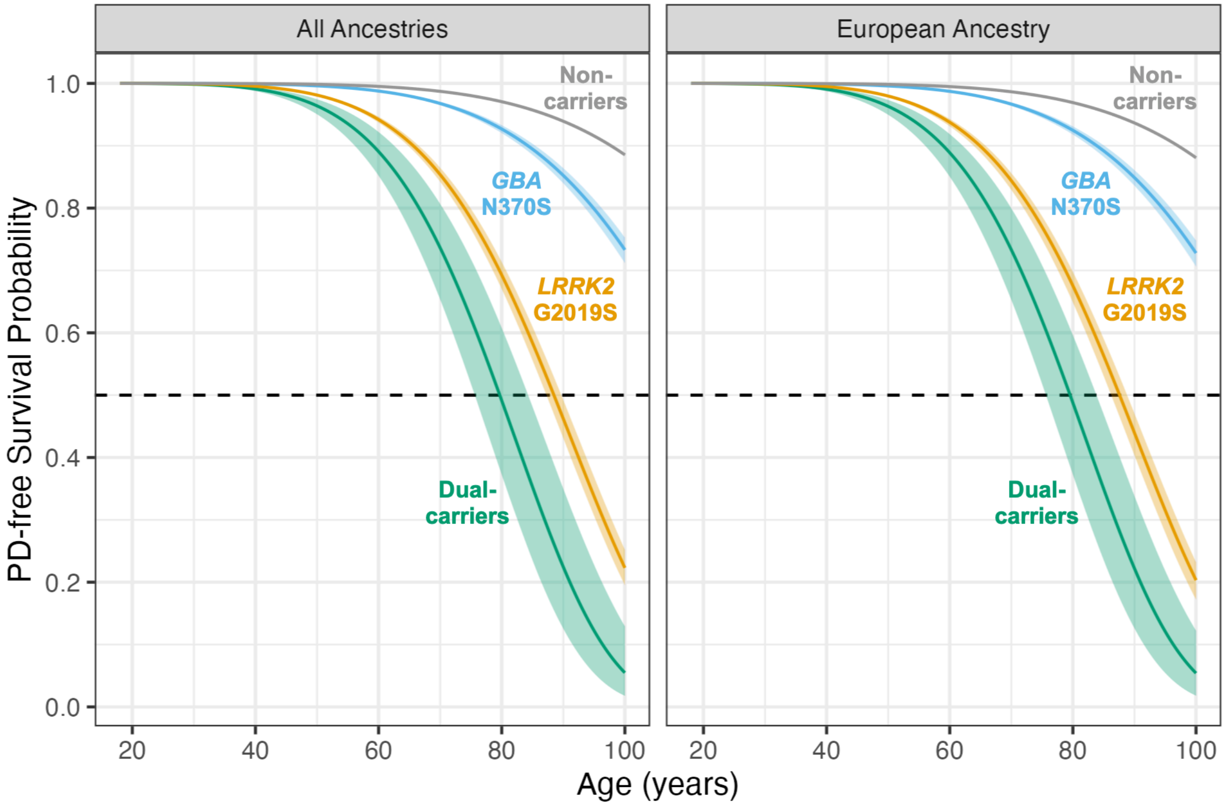


**eFigure 2. Predicted PD-free survival probabilities from the simple accelerated failure time models for participants across all ancestries (left) and participants with European ancestry (right).** The dashed line represents 50% survival probability for reference and shading denotes 95% confidence intervals. PD=Parkinson's disease.

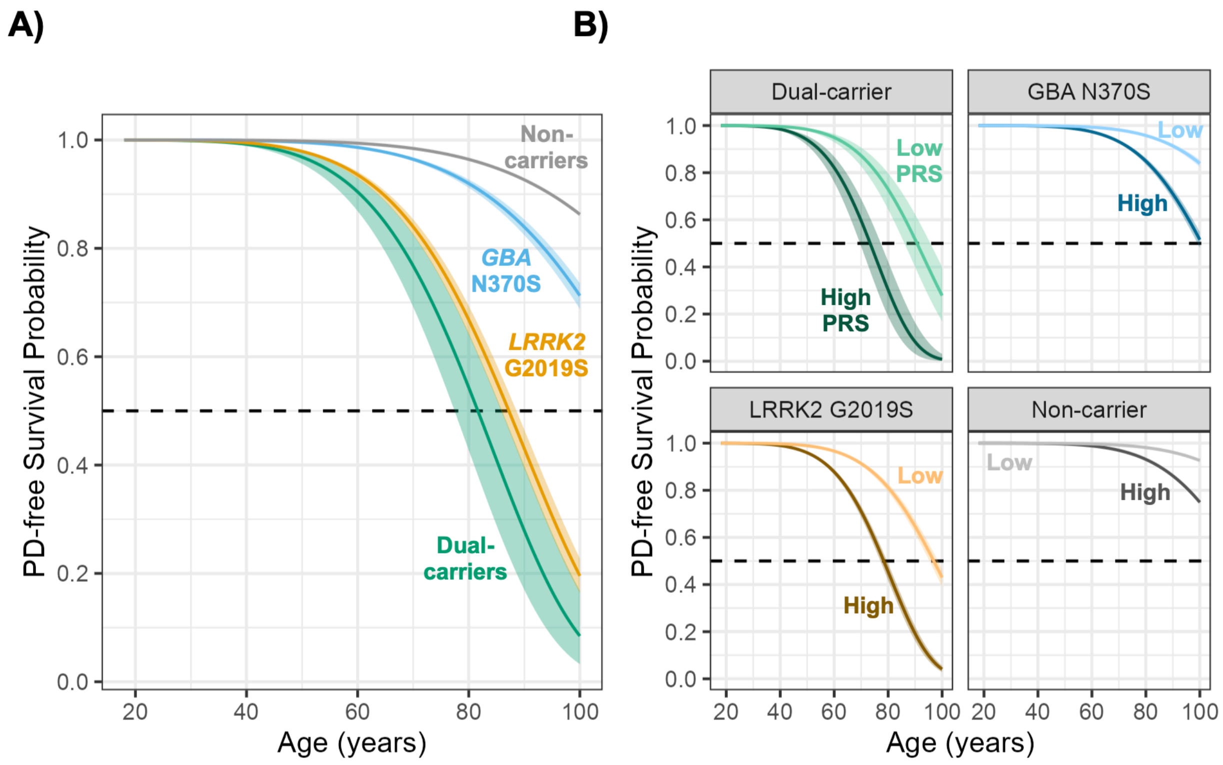


**eFigure 3. Predicted PD-free survival probabilities across different levels of PRS across European participants.** In both panels, curves represent predicted PD free survival probability for males using the sample means for 5 European ancestry PCs. The sample mean PRS was used for prediction in panel A, while low (10%) versus high (90%) PRS was used for the prediction in panel B. Shading denotes 95% CI.

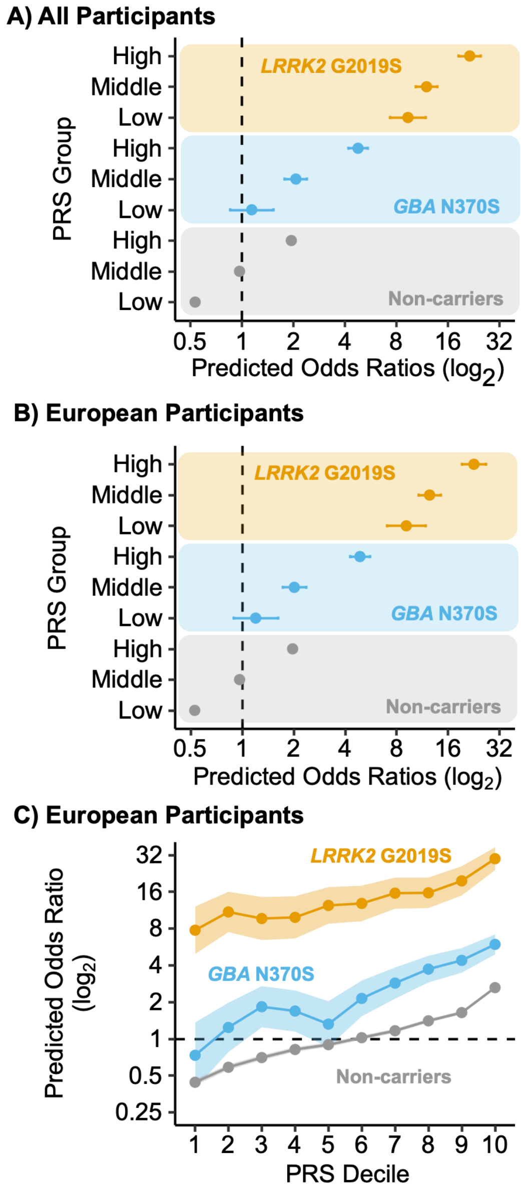


**eFigure 4. Risk of PD is stratified according to both carrier status and PRS.** A) PRS groups were defined according to PRS quantile with low PRS defined as bottom 25%, middle between 25-75%, and high as upper 25%. Predicted odds ratios reflect males with sample means for each ancestry principal component and mean age at study entry for all participants. Non-carriers with middle PRS served as the reference group. Error bars are 95% CIs. B) Similar results were observed when excluding non-European participants. Error bars are 95% CIs. C) PRS groups were defined according to PRS decile. Non-europeans were excluded. Predicted odds ratios reflect males with sample means for each ancestry principal component and mean age at study entry. Non-carriers at the 5th decile of PRS served as the reference group. Error shading reflects 95% CIs.

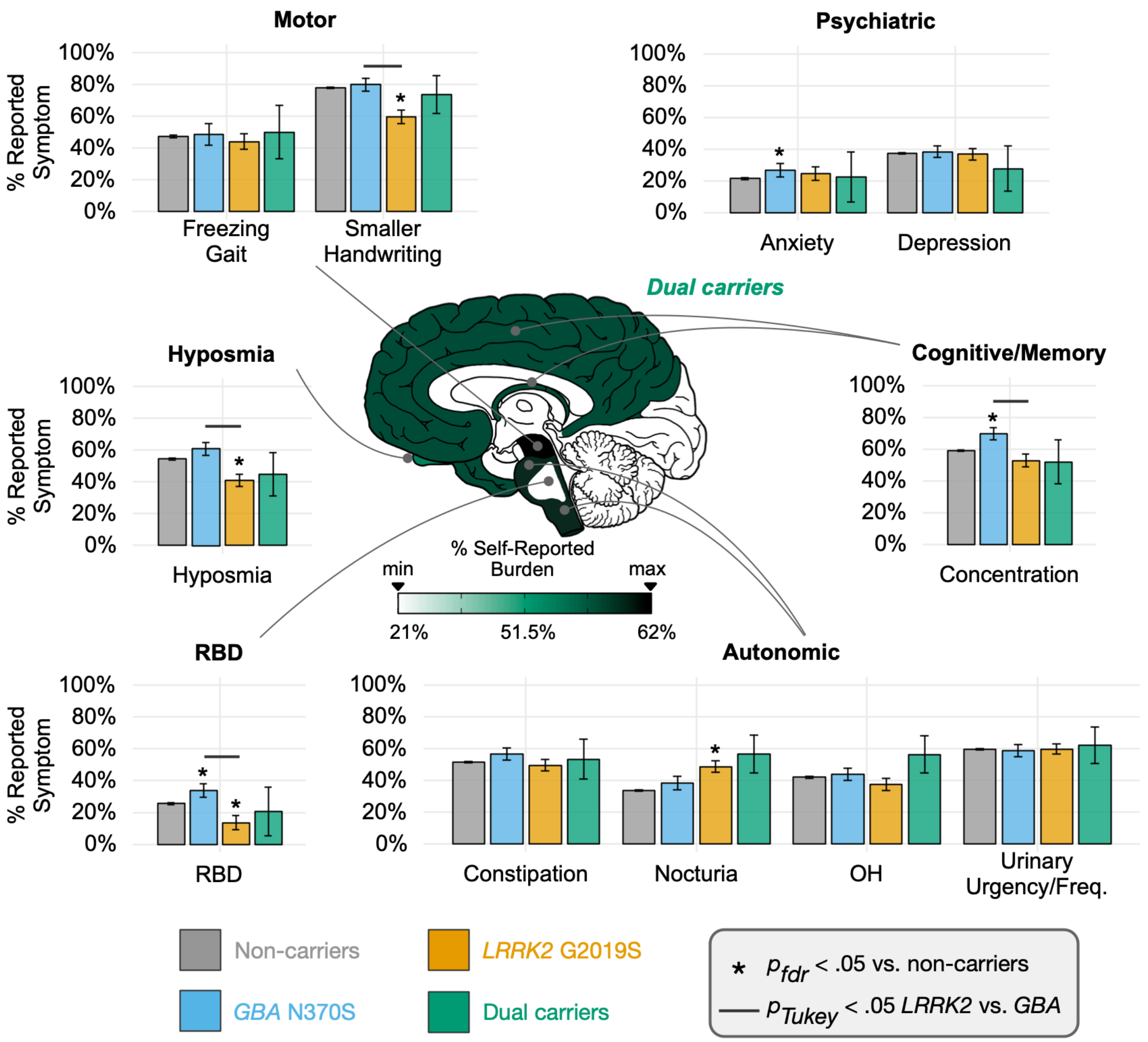


**eFigure 5. Self-reported symptom prevalence in *LRRK2* G2019S carriers with PD, *GBA* N370S carriers with PD, and non-carriers with PD (idiopathic PD), and dual *LRRK2* G2019S and *GBA* N370S carriers with PD.** Symptoms across questionnaires were aggregated into six domains and brain regions were shaded in approximated neuroanatomical regions according to average reported symptom burden across symptoms within domain: motor (substantia nigra), autonomic (brain stem excluding regions of the pons), cognitive/memory (cerebral cortex and limbic areas), hyposmia (olfactory bulb), REM sleep behavior disorder (RBD; areas of the pons), and psychiatric (no brain regions were shaded). The false discovery rate (FDR) was adjusted in carrier group comparisons to non-carriers within symptom domains. Comparisons between *LRRK2* G2019S, *GBA* N370S, and dual carriers were adjusted with Tukey's honestly significant difference tests. Error bars are *SE*. Descriptive statistics are not reported for several measures due to 23andMe data privacy policies ($n$<5).

A meta-analysis of genome-wide association studies. *The Lancet Neurology*, *18*(12), 1091–1102. https://doi.org/10.1016/S1474-4422(19)30320-5
